# Intravenous Iron Decreases Rehospitalizations but Doesn’t Change mortality in Patients Admitted with Acute Heart Failure and Iron Deficiency: A Systematic review and Meta-analysis

**DOI:** 10.1101/2021.07.11.21260344

**Authors:** Nischit Baral, Nabin R. Karki, Imran Akram, Ashiya Khan, Govinda Adhikari, Rohit Rauniyar, Basel Abdelazeem, Santosh K. Dhungana, Bandana Ranabhat, Arvind Kunadi

## Abstract

**Introduction:** The role of intravenous (IV) iron in chronic heart failure has been well studied, however, its role in acute heart failure (AHF) is less well-known. Including the recent AFFIRM-HF trial, we performed a systematic review and meta-analysis to highlight the role of IV iron in AHF with iron deficiency.

**Hypothesis:** We hypothesized that IV iron doesn’t change mortality or heart failure re-hospitalization rates in patients with AHF with iron deficiency.

**Methods:** We conducted a systematic review and meta-analysis of randomized controlled trials (RCTs) and cohort studies published from inception till June 30, 2021. We searched PubMed, MEDLINE, EMBASE (embase.com), and Cochrane database including only RCTs and Cohort studies. We also included one prospective and one retrospective Cohort studies and two RCTs in our meta-analysis. Eligible studies included adults with AHF, left ventricular ejection fraction less than 40%-50%, and able to receive IV iron therapy. Outcomes included re-hospitalization rates and overall mortality from 30 days to 52 weeks post randomization (in one RCT). We used random-effects model calculating Risk Ratio (RR) with 95% confidence interval (95% CI) using Review Manger 5.4 software. I^2^statistics was used to assess heterogeneity.

**Results:** There were total 1561 participants in both groups (IV iron and placebo/control) of four studies. The controls were comparable in both cohort studies and both the RCTs were well matched. Our results showed re-hospitalization in 278 of 833 (33.37%) patients in the IV iron/exposure group and 337 of 728 (0.46%) patients in the placebo/control group. The pooled result showed that the risk of rehospitalization was comparable across both groups (RR 0.85, 95%CI 0.62-1.17; I^2^=45%, P=0.14). However, subgroup analysis, including RCTs only showed that IV iron decreases re-hospitalization rate by 28% compared to placebo (RR 0.72, 95% CI: 0.64, 0.82, I^2^=0%, P<0.00001) but didn’t improve mortality when compared to placebo (RR 0.97, 95% CI: 0.73, 1.30, I^2^ =0%).

**Conclusions:** IV iron showed significant improvement in re-hospitalization rate for AHF hospitalizations in iron deficient patients but didn’t improve overall mortality. We need larger RCTs to further validate its effect on mortality.

## 1 Introduction

Iron deficiency with or without anemia is present in up to 50% of chronic heart failure patients [1]. Iron deficiency but not anemia may be associated with increased mortality in this population. Oral iron (ferrous salts) are poorly absorbed, requires prolonged use (> 6 months) and side effects (up to 70%) are common [2]. Intravenous (IV) iron replenishment offers rapid correction of iron deficiency and is safe. Iron therapy, mainly intravenous iron increases cardiac function, reduce hospitalizations, improve quality of life and decrease levels of negative markers such as NT-pro-BNP and CRP in patients with chronic heart failure. However, the role of intravenous iron in acute heart failure (AHF) is less well defined. With the recent publication of ground breaking AFFIRM-HF trial on intravenous iron in acute heart failure, there is excitement about its usefulness in acute decompensated heart failure as well [3]. However, still there is not as much supporting literature specific to the role of intravenous iron in acute heart failure. Iron replenishment is vastly underutilized in patients with iron deficiency hospitalized for acute decompensated heart failure [4]. An up-to-date systematic review about intravenous iron on acute heart failure and its beneficial effects on preventing hospitalizations may convince clinicians to use this modality more often. So, we performed a systematic review and meta-analysis to find the effects of intravenous iron on iron deficient patient with AHF.

## 2 Objective

To identify whether IV iron therapy reduces mortality or heart failure re-hospitalization rates in patients with AHF with iron deficiency.

## 3 Methods

### 3.1 Eligibility Criteria

We included randomized control trials (RCT) and cohort studies in English language, studies reporting patients with

1. Age more than 18 years or older
2. Patients hospitalized for acute heart failure
3. Patients having concomitant iron deficiency as documented by ferritin < 100 ng/mL or 100-299 ng/mL with transferrin saturation < 20%.

We excluded studies

1. On the role of IV iron in Chronic heart failure
2. Case report, case series or observational study
3. Non comparative study between IV iron and placebo
4. Studies done on oral iron

### 3.2 Study Design

The Preferred Reporting Items for Systematic Reviews and Meta-Analyses (PRISMA) statement for reporting systematic reviews as recommended by the Cochrane Collaboration was followed in this systematic review [5].

### 3.3 Information sources and Search Strategy

A systematic literature review using MEDLINE, Embase, Cochrane central, and clinicaltrials.gov was performed using the terms “Intravenous Iron”, “Ferric Carboxymaltose”, “Iron Deficiency”, “Acute Heart Failure”, “Acute decompensated heart failure”, “Randomized Controlled Trial”, and “Cohort studies” from literature published from inception till June 30, 2021. We limited our search to studies conducted in humans. Search was restricted to English language. We searched the reference section of included articles for additional reports.

### 3.4 Data collection, items, and outcome measure

Two reviewers independently performed the title and abstract screening and full text screening. Conflicts were resolved through consensus. Third reviewer was used whenever needed to resolve the conflicts. Our primary outcome of interest is rehospitalization rate for heart failure defined as hospital. We also looked into all-cause mortality as secondary outcome of interest.

### 3.5 Methodical quality assessment

We assessed the methodical rigor of the included studies using the modified Downs and Black checklist for RCTs and non-randomized studies [6]. The checklist has 27 items with a total possible score of 28. Papers were rated excellent if they scored above 25, good if they scored between 20 and 25, fair if they scored between 15 and 19 and poor if they scored <15. Each study was assessed by two independent investigators and discrepancies in scoring were resolved using consensus.

### 3.6 Publication Bias

We have only four studies included in the meta-analysis, so we did not construct a funnel plot for publication bias.

## 4 Results

### 4.1 Study selection

We identified 122 articles from rom PubMed/MEDLINE and 3 articles from Embase and Web of Science (Figure 1). Duplicate studies were removed by the software. We identified 9 studies. Finally, 9 articles were fully read and 5 articles removed for not meeting eligibility. Final qualitative and quantitative analysis was done with 4 studies (Figure 1).

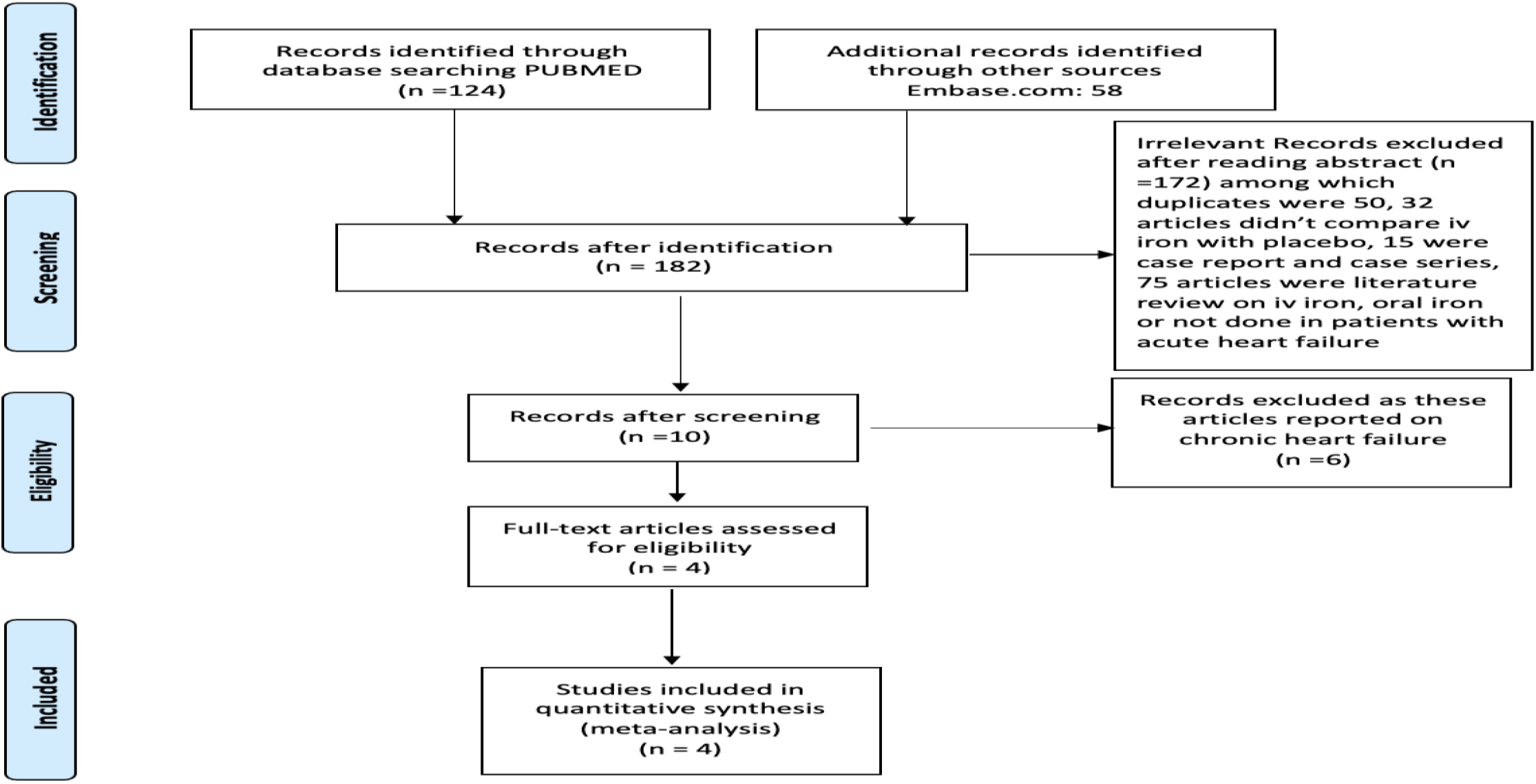

#### 4.1.1 Quality of included studies

Based on the Downs and black questionnaire tool, the quality of one of our studies was fair, one was excellent, and two were good. Table 1.

**Table 1:**
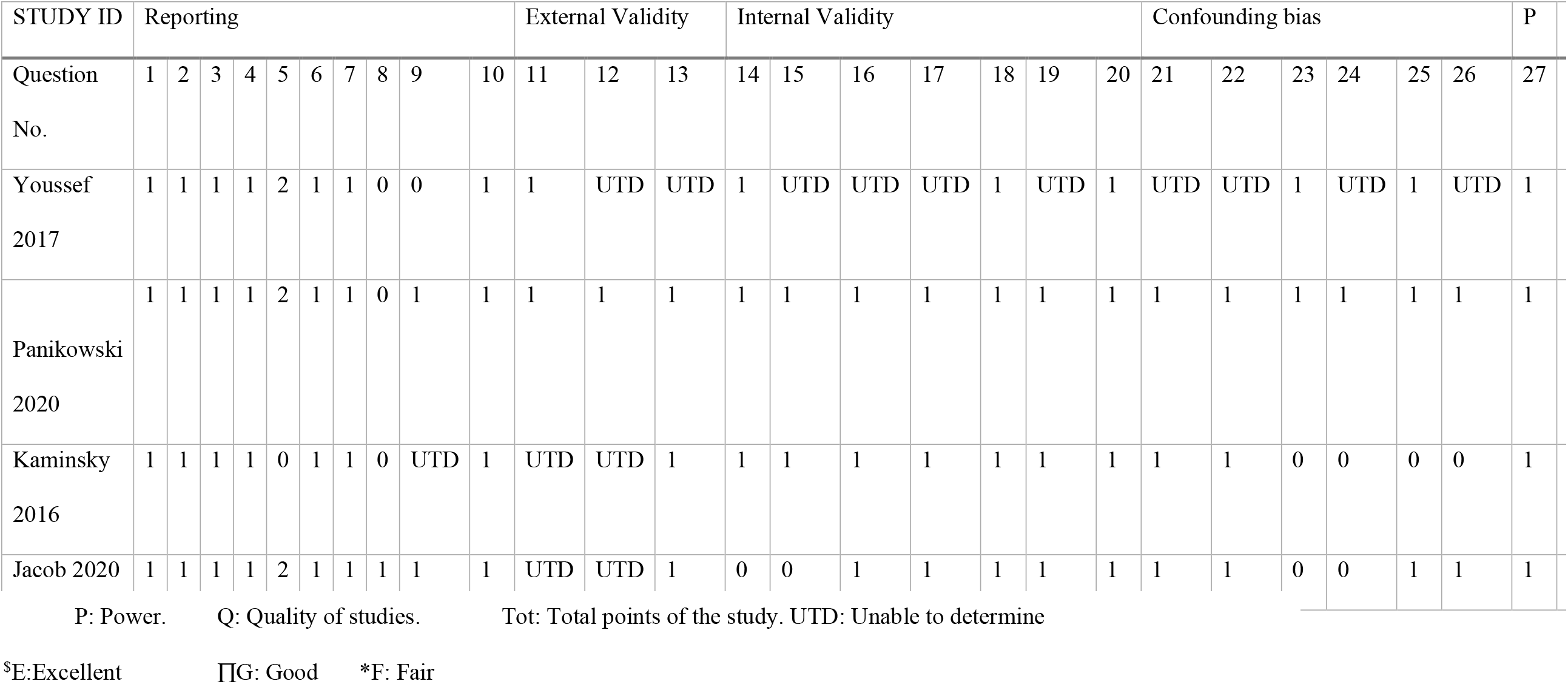
Quality of Included Studies

#### 4.1.2 Baseline Characteristics of Studies and follow up

We have included the baseline characteristics of the four included studies as follows. Table 2

**Table 2:** Baseline Characteristics of Included Studies

| STUDY | DESIGN | LOCATI<br>ON | INCLUSION | PARTICIPANTS | DU<br>RAT<br>ION | Outcome/Endpoi<br>nt: | Interve<br>ntion |
| --- | --- | --- | --- | --- | --- | --- | --- |
| Youssef<br>2017 | Randomized<br>Controlled Trial | Egypt | ADHF with NYHA class III-IV, LVEF < 40%, Hb < 13 g/dl in males, Hb < 12 g/dl in females, iron deficiency (ferritin < 100 mcg/L, or 100-299 mcg/L if transferrin saturation less than 20%) | 60, iv iron: 40, iv saline: 20 | 3<br>mont<br>hs | Rehospitalizatio<br>n and mortality<br>rates | Iv<br>Sucrose |
| Ponikowski<br>2020 | Multicenter<br>Randomized<br>Double Blind<br>Controlled Trial | Multinati<br>onal (45<br>nations) | Age > 18 years, Signs and symptoms of Acute Heart Failure, Elevated Natriuretic peptide and treated with at least 40 mg iv Lasix, LVEF < 50%, iron deficiency ferritin < 100 mcg/L, or 100-299 mcg/L if transferrin saturation less than 20%) | 1132, iv iron: 567, placebo: 565 | 52<br>wee<br>ks | Composite of<br>total heart failure<br>rehospitalization<br>and CV deaths | Iv<br>Ferric<br>carboxy<br>maltose |
| Jacob 2020 | Multicenter,<br>analytical,<br>prospective<br>follow-up<br>Cohort study | Spain | Patients with Acute heart failure (diagnosed as per ESC) requiring hospitalization. Our analysis included AHF and iron deficiency ferritin < 100 mcg/L, or 100-299 mcg/L if transferrin saturation less than 20%), | 191 iron deficient<br>group | one<br>year | In-hospital<br>mortality and 30-day readmission | Iv<br>Ferric<br>Carbox<br>ymaltos<br>e |
| Kaminisky<br>2016 | Retrospective<br>Cohort Analysis | USA | Age ≥ 18, Acute heart failure as primary discharge diagnosis, Hb < 13 g/dl, Transferrin saturation < 20% during admission, | 172, iv iron: 44,<br>control: 128 | 4<br>year<br>s and<br>8<br>mont<br>hs | Change in Hb<br>within 30 days of<br>therapy, 30-days<br>readmission | IV Iron<br>dextran<br>and IV<br>iron<br>sucrose |

### 4.2 Statistical Analysis

Outcomes from the individual studies were aggregated with Review Manager (version 5.4, Cochrane Collaboration, Oxford, United Kingdom) applying the Mantel-Haenszel test. Hazard ratio (HR) and 95% confidence intervals (CIs) were estimated using a random-effects method to account for the presence of variability among the studies. I^2^ statistic was used to assess heterogeneity. Two-tailed p-values <0.05 were considered to indicate statistical significance.

### 4.3 Heart Failure Hospitalization

Pooled data from four studies showed the incidence of heart failure rehospitalization in patients in the iv-iron group as 278 out of 749 patients and 337 of 728 patients in the placebo group. The pooled result showed that there was significant reduction in rehospitalizations in patients taking IV iron versus placebo (RR 0.88; 95% CI, 0.78, 0.99; heterogeneity I^2^ = 16%, p = 0.31). (Figure 2)

**Figure 2:**
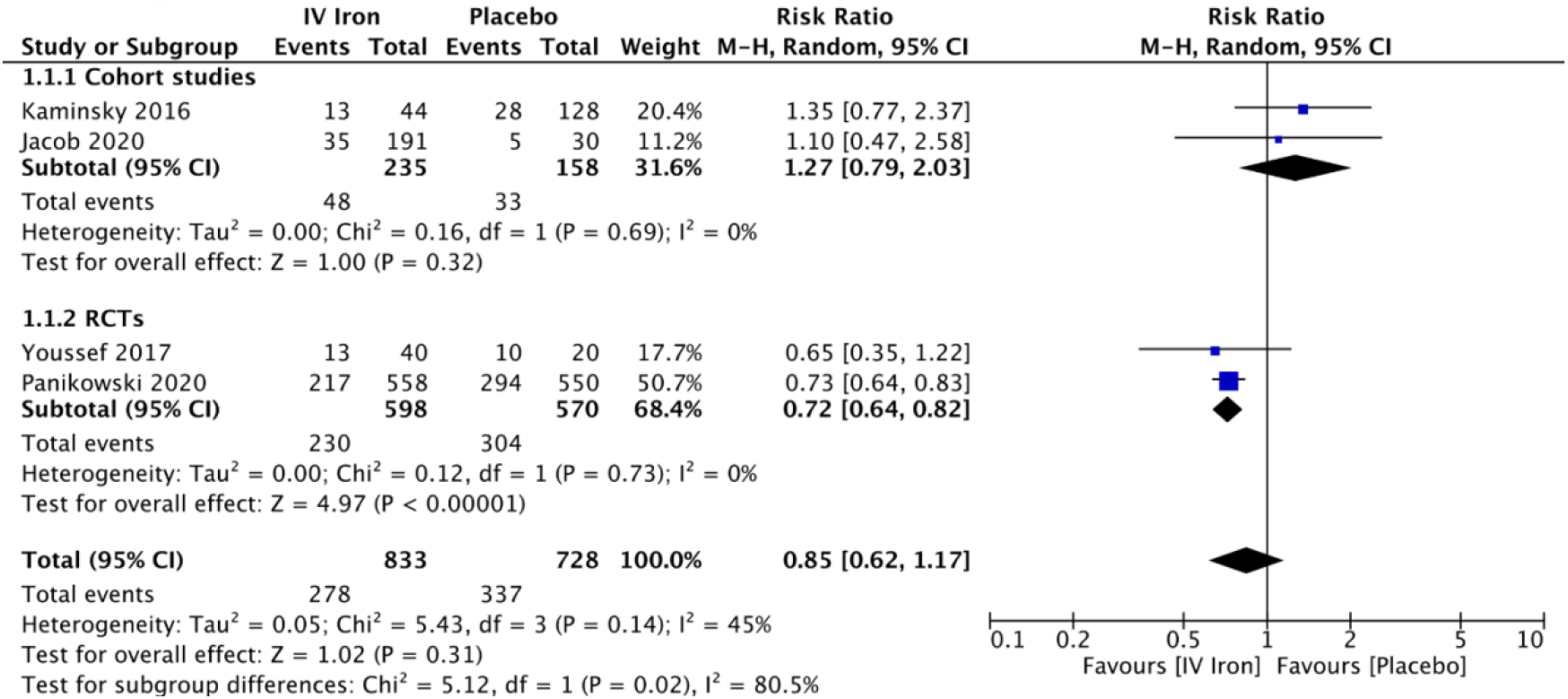
Forest plot showing the pooled RR of IV iron against placebo.

### 4.4 All-cause mortality

Pooled data from two studies showed that all-cause mortality is comparable between the iv-iron group and placebo group (HR 0.96; 95% CI, 0.72, 1.28; heterogeneity I^2^ = 0%, p = 0.95). (Figure 3)

**Figure 3:**
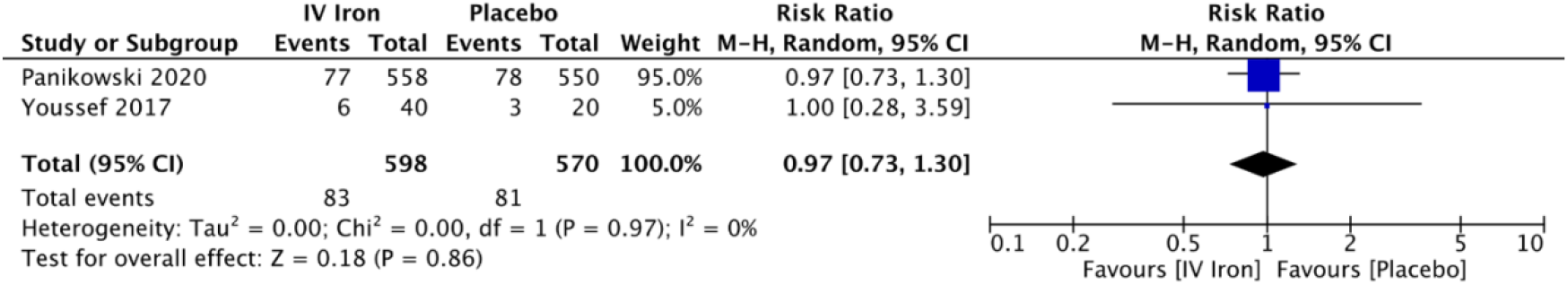
Pooled data of all-cause mortality in IV iron versus placebo

## 5 Discussion

Interest in treatment of anemia in heart failure states with intravenous iron was present as early as 1990s [7]. However, the earlier studies frequently used intravenous iron along with erythropoietin formulations and the results were disappointing [8]. When intravenous iron was used alone for treatment of anemia in patients with chronic heart failure, improvements in symptoms, exercise capacity and quality of life was noted [9][10]. This heralded an avalanche of studies on the subject.

Anemia in heart failure is multifactorial with contributions from inflammation, hemodilution, iron deficiency and use of ACE inhibitors. Interestingly, while inflammatory cytokines play a role, hepcidin only plays a minor role [11]. Most heart failure trials use ferritin <100 ng/mL or ferritin 100-299 ng/mL with transferrin saturation (Tsat) <20% to diagnose iron deficiency. These parameters are different from normal individuals but similar to other chronic inflammatory states. Screening for iron deficiency is class IC (evidence of benefit based on consensus or weaker evidence base) recommendation in European Society of Cardiology heart failure guidelines [12].In adults hospitalized with acute decompensated heart failure, iron deficiency was present in 68.6% among males and 75.3% among females (including 40.6% and 48.6% with absolute iron deficiency, respectively [4]. This number is about 12-15-fold higher than the general population of similar age group. Absolute iron deficiency is associated with risk of early readmissions [13]. However, only 9% patients received iron replenishment (oral, 8.7%; intravenous 0.2%) at admission [4]. Female sex, advanced age, higher level of NT-proBNP and CRP are associated with iron deficiency in heart failure states [14]. Iron deficiency also predicts poor exercise capacity and survival.

Older oral iron (ferrous salt) is poorly absorbed and have significant gastro-intestinal side effects. In chronic diseases, intravenous route is more efficient for iron replenishment because it bypasses malfunctioning intestinal barrier and make iron readily available to hematopoietic elements [15]. In IRONOUT HF, oral iron failed to show improvement in exercise capacity in patients with systolic heart failure [16]. In another small study with <20 patients that compared oral and intravenous iron, intravenous iron was superior in improving functional capacity of heart failure patients although both corrected anemia to similar extent [17]. Multiple meta-analyses have confirmed improvement in quality of life, increased 6-minute walk distance, reduction in hospitalization (± death) and symptom burden with intravenous iron in chronic heart failure [18][19][20]. Importantly, this is achieved with acceptable safety profile. However, iron therapy does not seem to affect mortality [21]. While upfront costs may seem high, intravenous iron is cost effective in the management of chronic heart failure [22][23]. Both American and European guidelines now support use of intravenous iron to improve functional status and quality of life in patients with chronic heart failure [24] [12].

AFFIRM-AHF study is the largest study exploring role of intravenous iron in acute systolic heart failure. It showed 26% relative reduction in hospitalizations (RR 0·74; 95% CI 0·58–0·94, p=0·013) with ferric carboxy-maltose. Intravenous iron was safe, with no effect on risk of cardiovascular death [3]. AFI-ID study included patients with heart failure with preserved ejection fraction [25]. All of the 3 studies by Jacob et al (AFI-ID), Kaminsky et al and Youssef et al reported non-significant trend towards reduced rehospitalization rates. However, neither of these studies were powered to detect differences in efficacy between groups [25][26][27]. The intravenous iron formulations (ferric carboxy-maltose, iron sucrose, iron dextran) all appear safe and efficacy is likely related to rise in hemoglobin. Intravenous iron supplementation confers 12% relative risk reduction in heart failure hospitalizations which is a clinically significant endpoint. Not surprisingly, no impact on mortality was seen, as with chronic heart failure. The cost-effectiveness of intravenous iron in acute heart failure remains to be studied.

The main limitation of our analyses is that our results are driven by the AFFIRM-AHF trial because of its relatively large sample size. Moreover, we did not consider the influences of type of iron formulation, dose and severity of heart failure due to incomplete individual patient level data. Another limitation of our meta-analysis is that the primary outcome of rehospitalization was followed up at different duration which can lead to different outcomes.

## 6 Conclusion

Intravenous iron has now proven benefit in preventing rehospitalization in AHF patients with concomitant iron deficiency. No mortality benefit is present though. Our study further supports these findings from multiple individual studies. We hope this synthesis of available evidence on the topic will result in clinicians and guidelines embracing it.

## Data Availability

Data is available in the main text itself.

